# Altitude as a protective factor from COVID-19

**DOI:** 10.1101/2020.08.03.20167262

**Authors:** Timothy M. Thomson, Fresia Casas, Harold Andre Guerrero, Rómulo Figueroa-Mujíca, Francisco C. Villafuerte, Claudia Machicado

**Affiliations:** Institute for Molecular Biology, National Science Council (IBMB-CSIC), Barcelona, Spain; Networked Center for Biomedical Research in Hepatic and Digestive Diseases (CIBER-EHD), Instituto Nacional de la Salud Carlos III, Madrid, Spain; Laboratory of Translational Research and Computational Biology, Facultad de Ciencias y Filosofía-LID, Universidad Peruana Cayetano Heredia, Lima, Perú; Laboratorio de Fisiología del Transporte de Oxígeno, Facultad de Ciencias y Filosofía-LID, Universidad Peruana Cayetano Heredia, Lima, Perú; Institute for Biocomputation and Physics of Complex Systems (BIFI), Zaragoza, Spain

**Keywords:** COVID-19, Peru, altitude, chronic hypoxia

## Abstract

The COVID-19 pandemic had a delayed onset in South America compared to Asia (outside of China), Europe or North America. In spite of the presumed time advantage for the implementation of preventive measures to help contain its spread, the pandemic in that region followed growth rates that paralleled, and currently exceed, those observed several weeks before in Europe. Indeed, in early August 2020, many countries in South and Central America presented among the highest rates in the world of COVID-19 confirmed cases and deaths per million inhabitants. Here, we have taken an ecological approach to describe the current state of the pandemic in Peru and its dynamics. Our analysis supports a protective effect of altitude from COVID-19 incidence and mortality. Further, we provide circumstantial evidence that internal migration through a specific land route is a significant factor progressively overriding the protection from COVID-19 afforded by high altitude. Finally, we show that protection by altitude is independent of poverty indexes and is inversely correlated with the prevalence in the population of risk factors associated with severe COVID-19, including hypertension and hypercholesterolemia. We discuss long-term multisystemic adaptive traits to hypobaric hypoxia as possible mechanisms that may explain the observed protective effect of high altitude from death due to COVID-19.

## Introduction

The expansion of the SARS-CoV-2 pandemic from Wuhan, China, has followed a rapid international spread in line with global travel patterns and slower, more complex within-country spread dynamics, attuned not only by internal migration patterns but also by a broad array of additional local determinants, including non-pharmaceutical policy interventions^1^, population density^2^, demographics^3^, health care system capacity and quality^4^, inequality and poverty^5^, age^6-8^, co-morbidities^8-10^, ethnicity^8,11^, genetic factors^12^, household structure^13^, air quality^14^ or climate^15,16^. In South America, subsequent to the first confirmed case of COVID-19 reported on February 25 in São Paulo, Brazil, the pandemic has followed an apparently protracted initial phase of spread, before entering an exponential growth phase of cases and deaths, becoming the latest global epicenter of the pandemic^17^. In some cases, in particular Brazil, failure to implement mandatory nation-wide non-medical mitigation measures, including the use of face masks^18^ and application of social distancing, may help explain the uncontrolled spread of the pandemic. However, in other countries, like Peru, introduction of several such measures at an early phase of the pandemic, albeit confusing as to the use of face masks, has not prevented its spread and high mortality rates in the country as a whole.

In part due to the relative scarcity of detailed data^19^ and to underreporting, few studies^20,21^ have explored the dynamics and specific characteristics of the COVID-19 pandemic spread in South America. Until recently^22^, limited availability of viral sequence information^23-25^ has also prevented genomic epidemiological approaches to understanding viral spread, variation and possible heterogeneous pathogenesis^26^ in specific geographical and population contexts in the region. In spite of these information shortcomings in the early stages of the pandemic in South America, accumulating data are making possible to conduct more detailed epidemiological analyses, essential to inform policies intended to protect the populations at higher risk. Here, we have analyzed open-access data for confirmed COVID-19 cases and deaths in Peru, by means of temporal and spatial analyses, along with association analyses with demographic parameters, socio-economic determinants and co-morbidities in an ecological approach to the pandemic. Our analysis reveals a significant protection from COVID-19 contagion and death conferred by high altitude, independent of population density, distance from the pandemic epicenter or poverty levels.

## Results

### Age distribution and female-to-male rates of COVID-19 in Peru

The COVID-19 pandemic in Peru has followed a dynamic similar to that of other South American countries. Unlike the majority of European countries or North America, in which multiple import events have seeded subsequent viral spread, the spread in South America can often be traced to a single import event per country^22^. In order to better understand the dynamics of the pandemic in Peru and potential ecological peculiarities, we have undertaken a systematic longitudinal and geographical analysis of all cases and deaths registered in Peru through July 17, 2020. (Table S1). General data on the pandemic in Peru are readily available to the public through several information channels (https://cs.ucsp.edu.pe/monitor-covid/; https://covid19.minsa.gob.pe/sala_situacional.asp). We dissected such coarse data into finer detail, by analyzing the age distribution and female-to-male ratios of cumulative cases diagnosed and deaths registered on four separate dates (April 10, May 15, June 19 and July 17) in each of the larger territorial divisions of the country (i.e., departments, each consisting of provinces, that encompass several districts). In these analyses, we have avoided the case-fatality ratio metric as it may obscure the geographical dynamics of the pandemic in the event of concomitant evolution of cases and fatalities^27^ and because of a degree of uncertainty in territorial differences in testing coverage. Importantly, all our comparative analyses are performed with normalized case and death numbers both for population density [(cases or deaths/(population/Km^2^)] and for total population (cases or deaths/10^6^ population).

While the age distribution of cases at the overall national level was stable along the entire study period, with a median age 42 on all four dates, similar to that reported for countries like China^28^, several departments showed a significant departure from the nationwide distribution, not only on the early cut-off point of April 10, but also on later dates (Fig. 1A). This was particularly evident in departments distant from the large coastal urban centers, such as Cajamarca, Cusco and Huancavelica (in the Andes) or San Martín, Amazonas and Madre de Dios (in the Amazon, or “selva”, region), with median ages for cases ranging from 27 (Madre de Dios; April 10 cumulative) to 39 (Madre de Dios; May 15, June 19 and July 17). These relatively young age distributions for COVID-19 cases are reminiscent of those for South Korea^28^, a reflection of generally younger demographics as compared to Italy, Spain or other European countries^29^. The nationwide age distributions of death from COVID-19 were also stable over time (mean age 66). The distribution of death from COVID-19 by department were variable on the earlier dates, likely owed to the low number of deaths in several departments early in the pandemic (Fig. 1A), but were generally uncoupled from age distribution of cases, as expected from the relatively low incidence of death from SARS-CoV-2 infection in the young^29^. Also as expected^7,9^, cases and, particularly, deaths, affected more men (0.72 to 0.74) than women nationwide (Fig. 1B). However, notable exceptions were several departments in which much of their territories are located at high altitude (> 2,500 m), namely Apurímac, Pasco and Puno, as well as Moquegua (featuring inhabited districts with altitudes ranging from sea level to above 4,000 m), where death from COVID-19 affected more women (0.51 to 0.82 for the June 19 or July 17 cut-offs) than men (Fig. 1B).

**Figure 1.**
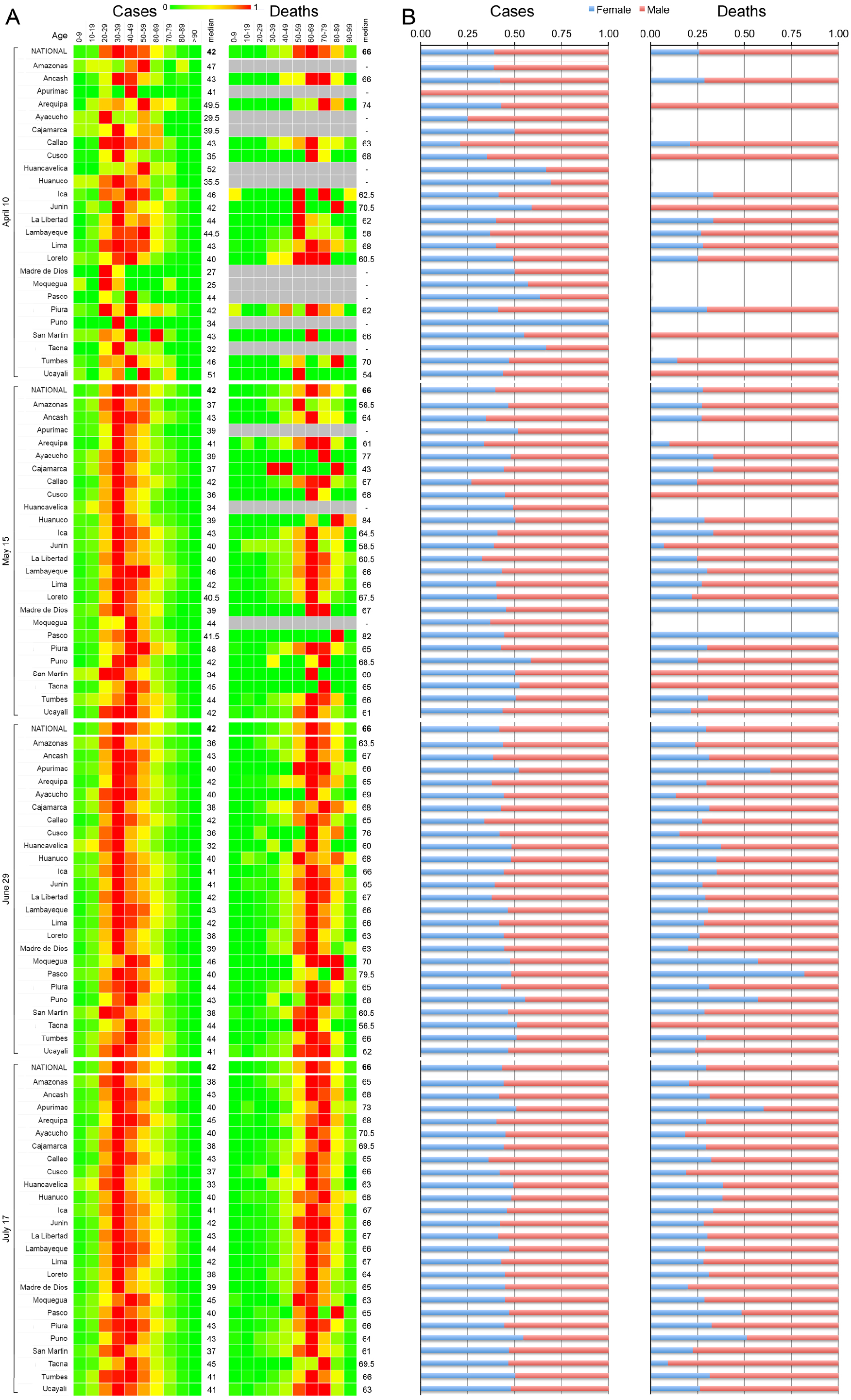
Longitudinal distribution of COVID-19 cases and deaths in Peru accumulated on four cut-off dates, by department, age range and sex. **A**. Heatmap of frequencies of distributions of cumulative cases and deaths at each of the indicated dates for the indicated age ranges and departments. Median ages for cases and deaths for each department and each cut-off date are shown to the right of the heatmaps. Nationwide heatmaps corresponding to age range distribution frequencies and median ages are also shown for each date. **B**. Histograms of relative frequencies of cumulative male and female cases and deaths on the indicated dates at each department (as indicated in panel A). Nationwide female:male relative frequencies are also shown for each date.

Peru presents a peculiar orographical arrangement, in which low-altitude regions are both on the coast and inland (Amazon or “selva” regions), separated by the high-altitude Andean range. This setting is uniquely suited to test hypotheses on the potential health impacts of altitude as a factor independent from other factors such as population density or distance from large urban centers. By geographically mapping population density-normalized case and death rates at the district level, we corroborated in more detail coarser publicly available maps, indicating that higher case and death rates generally corresponded to coastal and Amazon (“selva”) districts, relatively sparing districts located in the Andes range (Fig. 2). Interestingly, although the recognized initial epicenter of the pandemic is attributed to the Lima region, which indeed presents the higher overall number of cases and deaths in Peru (Table S1), the population density-normalized data indicated an early, higher intensity epidemic in relatively distant coastal districts located North and South of Lima, as well as in a number of Amazon districts. As the pandemic progressed, the case and fatality rates progressively increased over time in a number of high-altitude districts. However, the majority of such districts were relatively unaffected, with case and death rates per population density below 1 (Fig. 2). Coastal districts are those with the largest urban centers and higher population densities. That many “selva” districts, geographically distant and not readily accessible from the coast, had case and death rates as high as many coastal districts suggests that proximity to large coastal urban centers alone is not a major factor explaining the high COVID-19 case and death rates.

**Figure 2.**
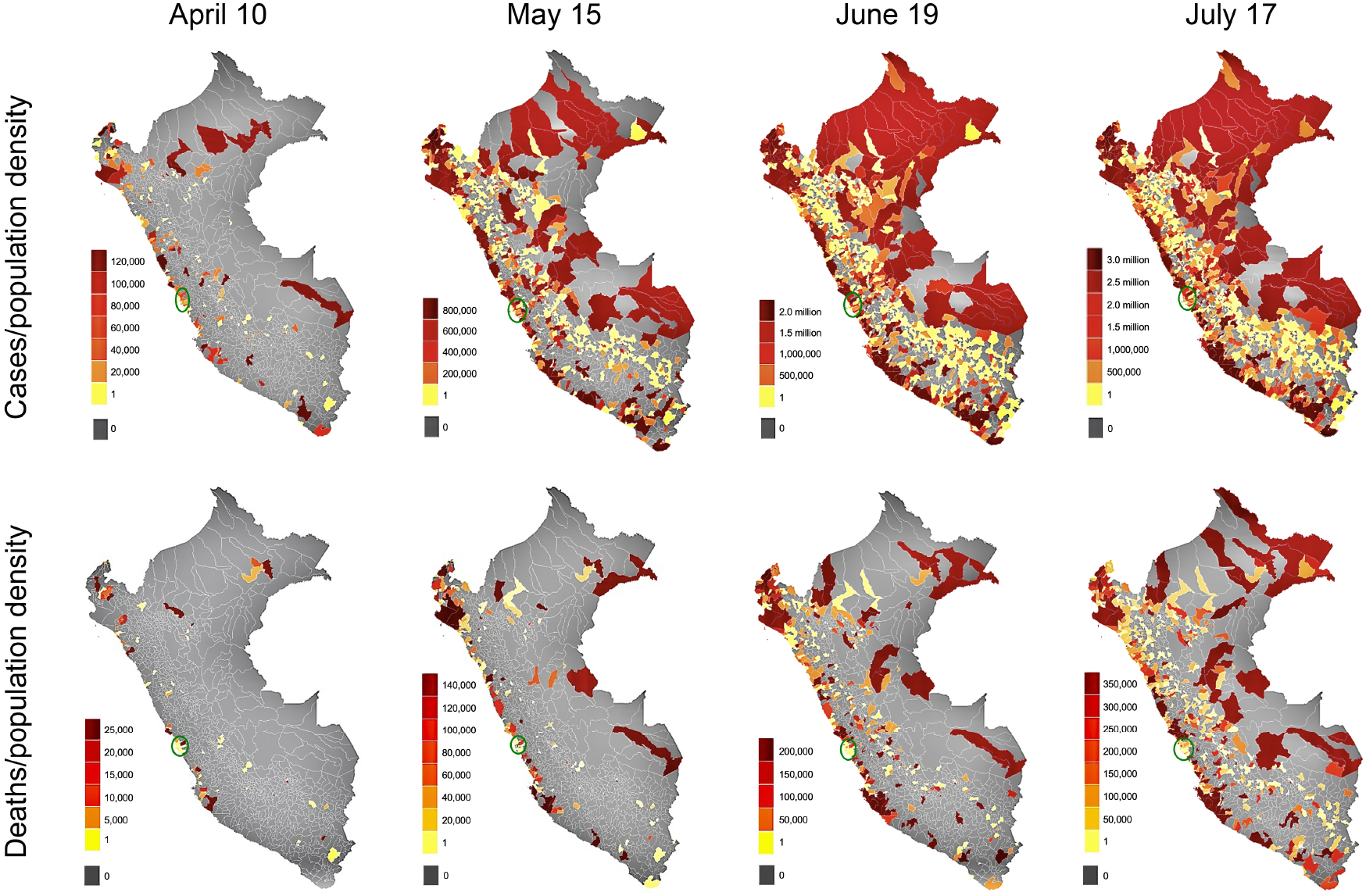
District-level map of case and death rates in Peru, normalized for population density. Cumulative COVID-19 case and death rates per population density [cases or deaths/(persons/Km^2^)] on each of the indicated cut-off dates are mapped at the district level. Heatmaps and their equivalencies are specific for each date. The numbers indicate the upper limits of value range. Blue circles are indicators of the Lima metropolitan region.

### Distribution and relative risk of COVID-19 case and death rates as a function of altitude

Given the above geographical distribution of COVID-19 cases and deaths, and following up several reports that have suggested an inverse correlation between altitude and COVID-19 cases and severity^30-32^, we analyzed the incidence of confirmed COVID-19 cases and deaths at various altitude intervals. High altitude is defined as terrain located above 2,500 m and thus we first used this altitude threshold to stratify our data. It can be seen (Fig. 3) that the rates of COVID-19 cases and deaths in districts at altitudes below 2,500 m were significantly higher than those at altitudes above this mark on all three cut-off dates analyzed. We further stratified populations into 1,000-m intervals, and found highly significant differences in case and death rates in comparisons between sites located below 1,000 m and all other sites located above 1,000 m on all three cut-off dates (Fig. 4). Smaller differences in case and death rates were observed in comparisons between altitude intervals above 1,000 m.

**Figure 3.**
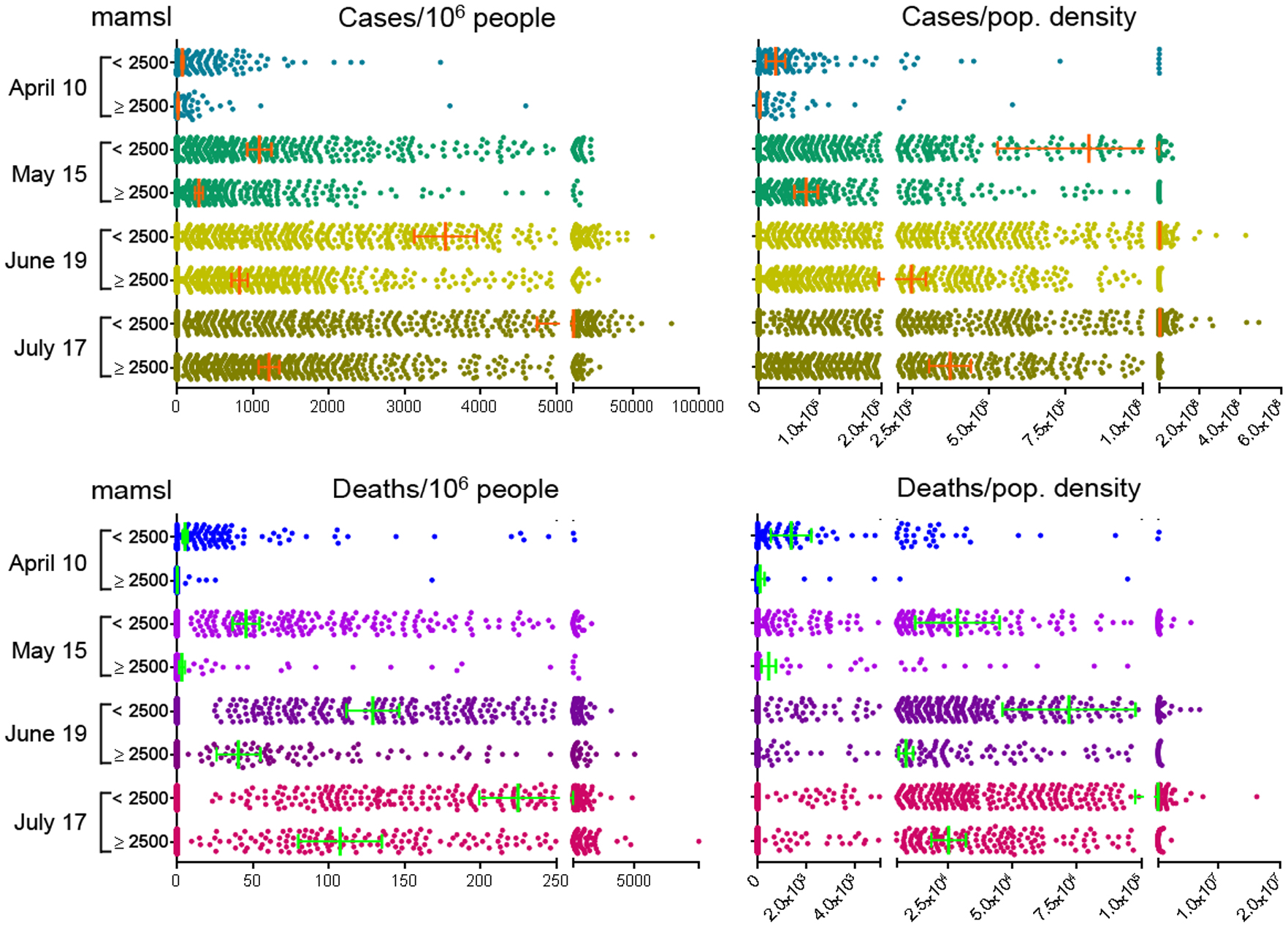
Distribution of case and death rates per million people or population density in districts located below or above 2,500 m of altitude. Cumulative deaths and cases per 10^6^ population or population density were plotted on each of the 4 indicated dates. Green and orange vertical lines denote mean values and horizontal lines 95% confidence intervals. All pairwise comparisons between low- and high-altitude strata on all 4 dates yielded P values < 0.0001 (Mann-Whitney nonparametric test). mamsl, meters above mean sea level.

**Figure 4.**
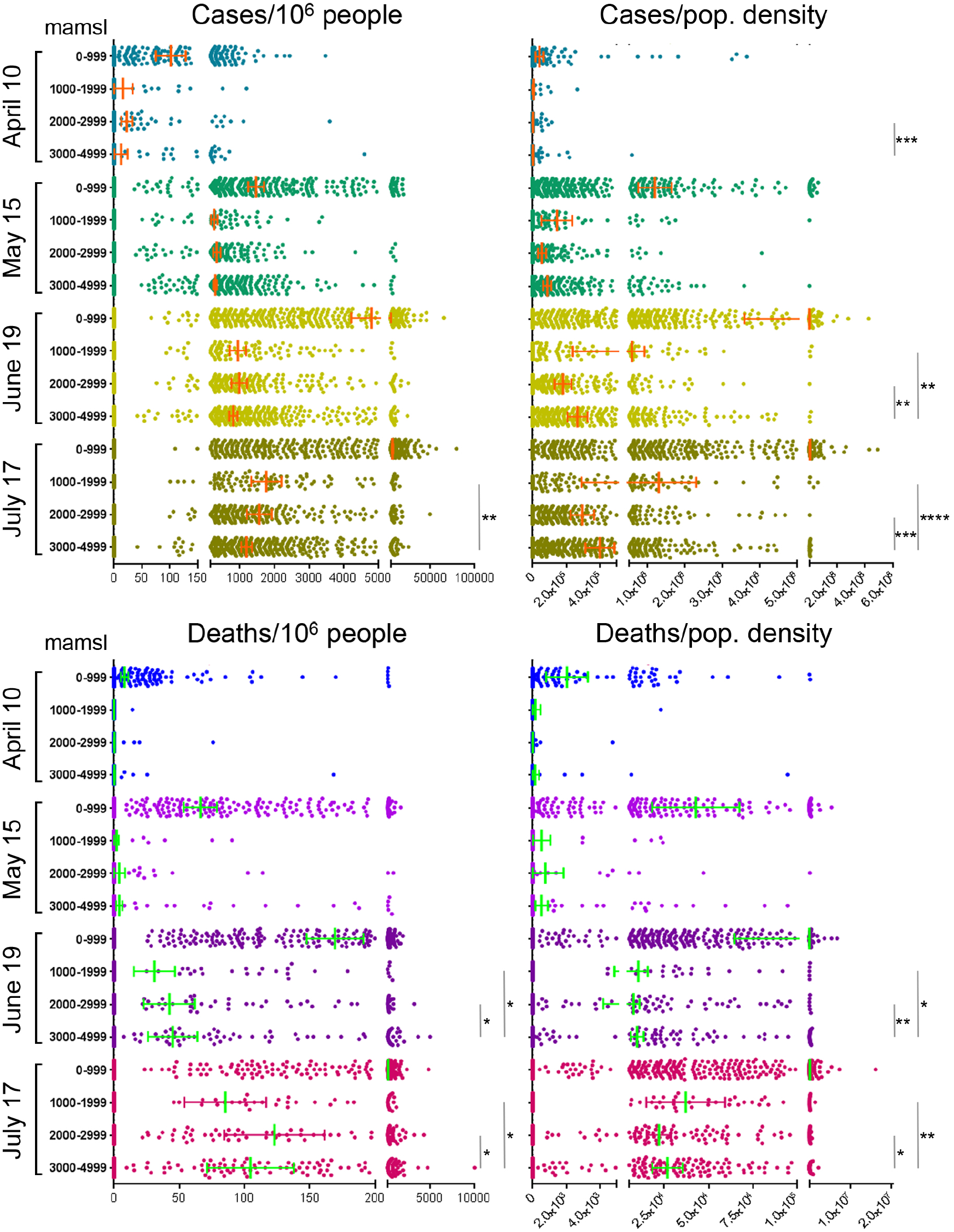
Distribution of case and death rates per million people or population density in districts stratified at 1,000 m intervals. Green and orange vertical lines denote mean values and horizontal lines 95% confidence intervals. P value ranges are are denoted as asterisks: *, < 0.05; **, < 0.001; ***, < 0.0001. (Mann-Whitney nonparametric test). Not shown are significant level indicators for comparisons between case or death rates between the 0-999 m interval and all other altitude intervals on all dates, all highly significant (< 0.0001). The *x*-axes were segmented for better visualization.

Consistently, relative risks (RR) were significantly higher for COVID-19 case and death rates in districts at altitudes lower than 2,500 m, with 11.4-fold (April 10 cut-off), 7.6-fold (May 15), 8.2-fold (June 19) and 6.5-fold (July 17) RR for case rates and 13.1-fold (April 10), 21.6-fold (May 15), 10.5-fold (June 19) and 6.5-fold (July 17) RR for death rates (Fig 5A, Table S2). Likewise, RR for case and death rates were significantly higher in comparisons between sites below 1,000 m and all other altitude intervals above 1,000 m, and generally not significant when comparing 1,000-m intervals above 1,000 m (Fig. 5B, Table S2). An exception to the latter was observed when comparing RR between the 1,000-1,999 m and 3,000-3,999 m intervals, which suggests small, but significant, relative protection from SARS-CoV-2 infection and COVID-19 death conferred by the higher altitude range (Fig 5B, Table S2). This suggests that altitudes above 1,000 m associate with a protective effect from COVID-19, and that altitudes above 1,000 m do not significantly add further protection. Of note, the potential protection conferred by altitude, although strong and consistently significant for all altitude ranges above 1,000 m as compared to those below 1,000 m, declined over time (Fig. 5B, Table S2). Interestingly, a progressively dwindling protection conferred by altitude over time was conspicuous in the 1,000-m interval comparisons (Fig. 5B). This might be attributed to either the apparent protection being an artifact related to testing coverage or data registry shortcomings, or to factors associated with the dynamics of the pandemic (see below).

**Figure 5.**
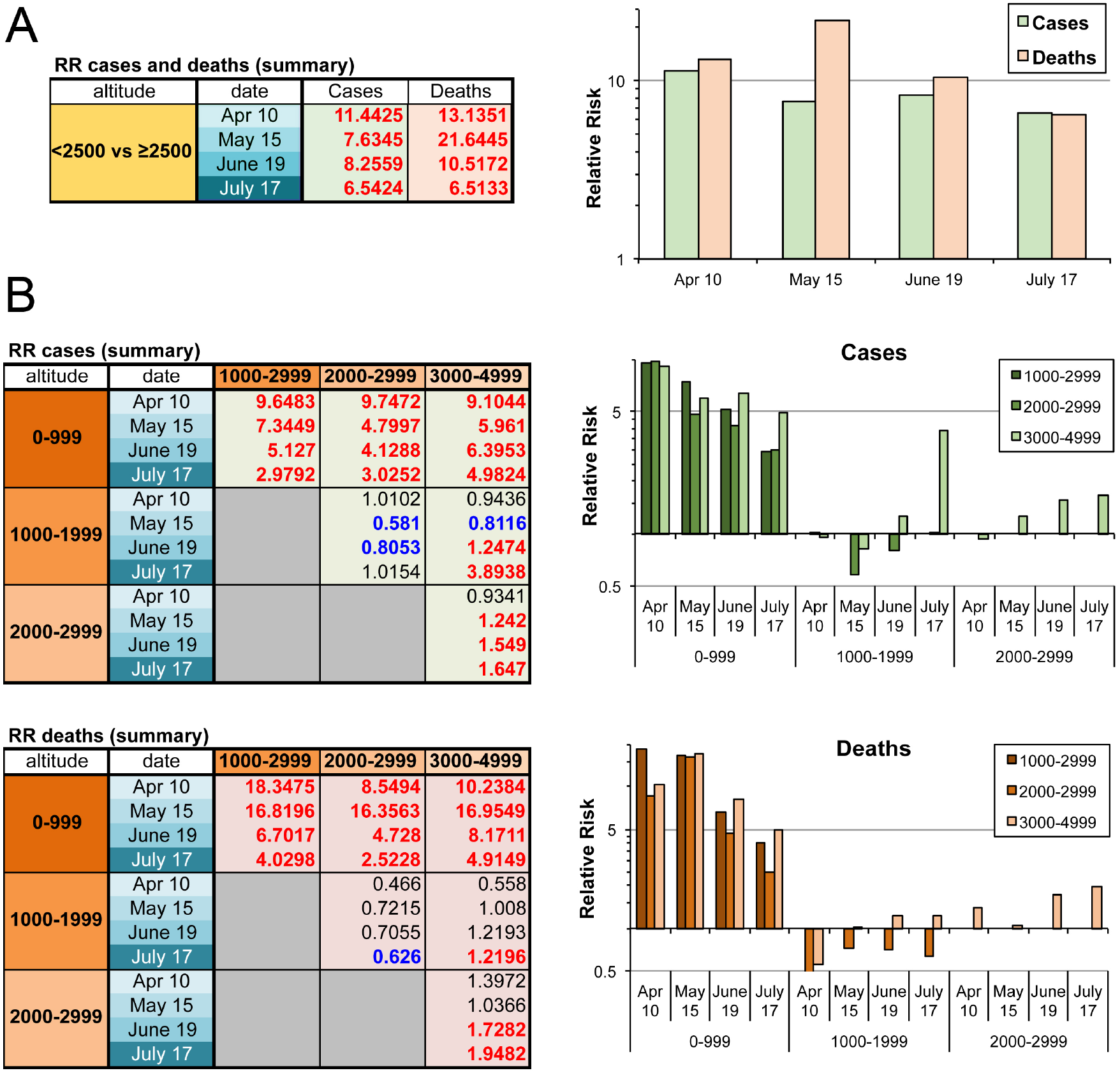
Relative risks for SARS-CoV-2 positiviy and death from COVID-19 in comparisons between low-altitude *vs*. high altitude districts. **A**. <u>Left panel</u>, table of relative risk values calculated for cumulative registered cases and deaths in districts below or above 2,500 m on the indicated dates. Given the uncertainty of the testing coverage, total populations in each altitude range were used as surrogate control groups in these calculations. <u>Right panel</u>, graphical representation of the values on the left table. **B**. <u>Left panels</u>, tables of relative risk values calculated for cumulative cases and deaths in districts at 1,000-m altitude intervals. Total populations in each altitude range were used as surrogate control groups. <u>Right panels</u>, graphical representations of the values on the left tables. Logarithmic scales were applied in the *y* axes for improved visualization of negative relative risk values. In all tables, values in bold are significant, red fonts denote increased relative risk for districts in the altitude range in the leftmost columns and blue fonts denote decreased relative risk in the corresponding pairwise comparisons Values in regular font denote non-significant relative risks. Confidendce intervals and p values are detailed in Table S2.

Because the apparently abrupt drop in death rates from districts below and above 1,000 meters of altitude might be an artifact of the stratification used, we stratified districts into 500-m altitude intervals and calculated the mean value of population density-adjusted death rates for each interval. We observed that the decline in death rates was indeed gradual, albet not linear, from the first altitude interval up to the boundary between the 1500-1999 and 2000-2499 m intervals, best visible in the July 17 cut-off date (Fig. 6). The low death rates at high altitude on earlier cut-off dates, close to or equal to zero, prevent an adequate visualization of this decline until later dates.

**Figure 6.**
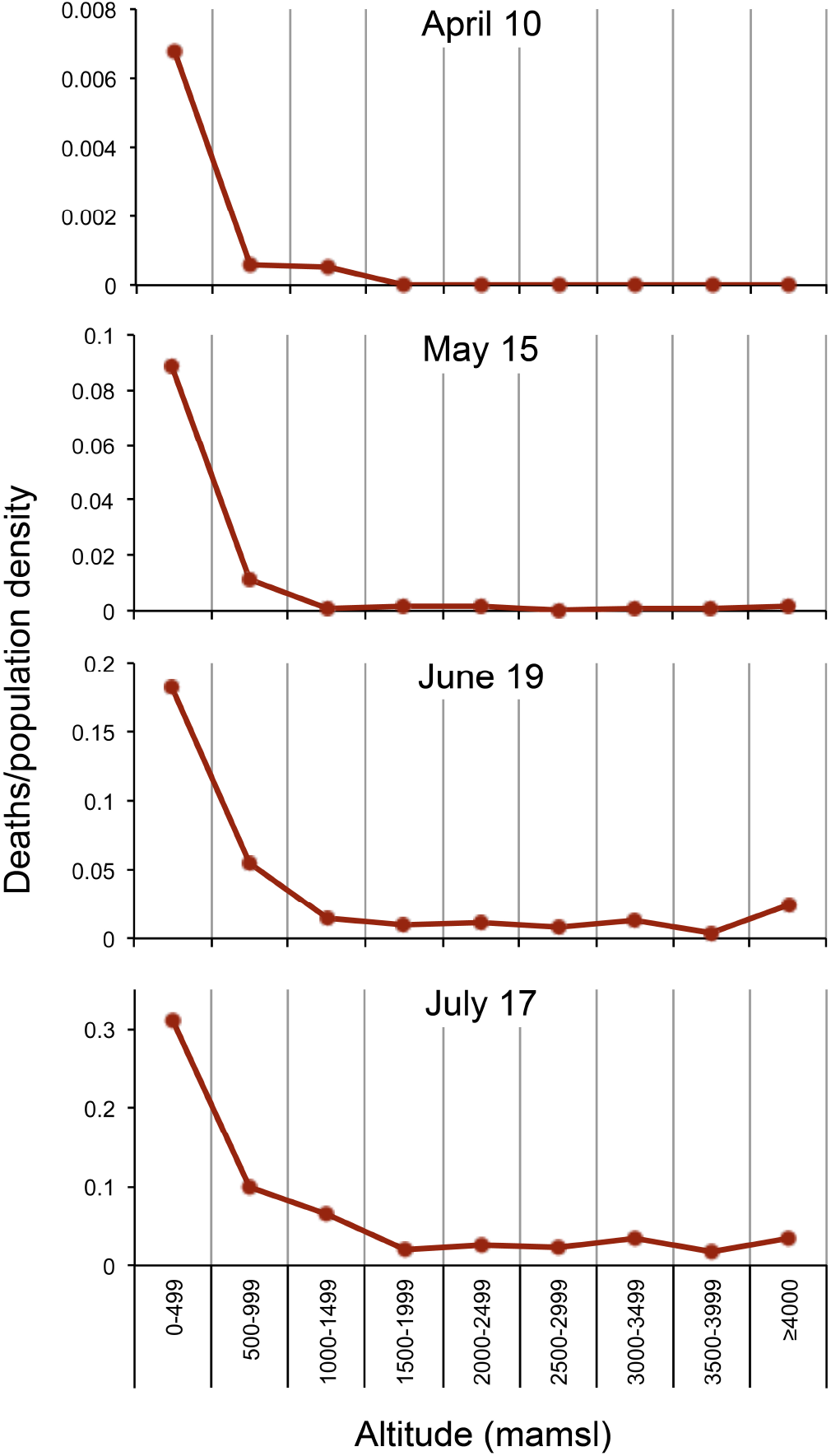
Gradual decline in COVID-19 death rates with increasing altitude. Population density-adjusted COVID-19 deaths (deaths/population density) determined for all districts were stratified into 500-m altitude intervals. Mean values were calculated and plotted for each atitude interval. mamsl, meters above mean sea level.

As crowding and high population density are established transmission factors for viral respiratory diseases, including COVID-19 (ref. 2), we further compared COVID-19 death rates between pairs of districts with equivalent population densities but located at different altitudes, either below 1,000 m (low altitude) or above 2,500 m (high altitude). With few exceptions, these pairwise comparisons yielded significantly greater death rates in districts at low altitude than at high altitude (Fig. 7). This was also observed in pairwise comparisons between low- and high-altitude districts within the same department (Fig. S1). Collectively, these observations suggest a protective effect from COVID-19 conferred by altitude, independent of population density.

**Figure 7.**
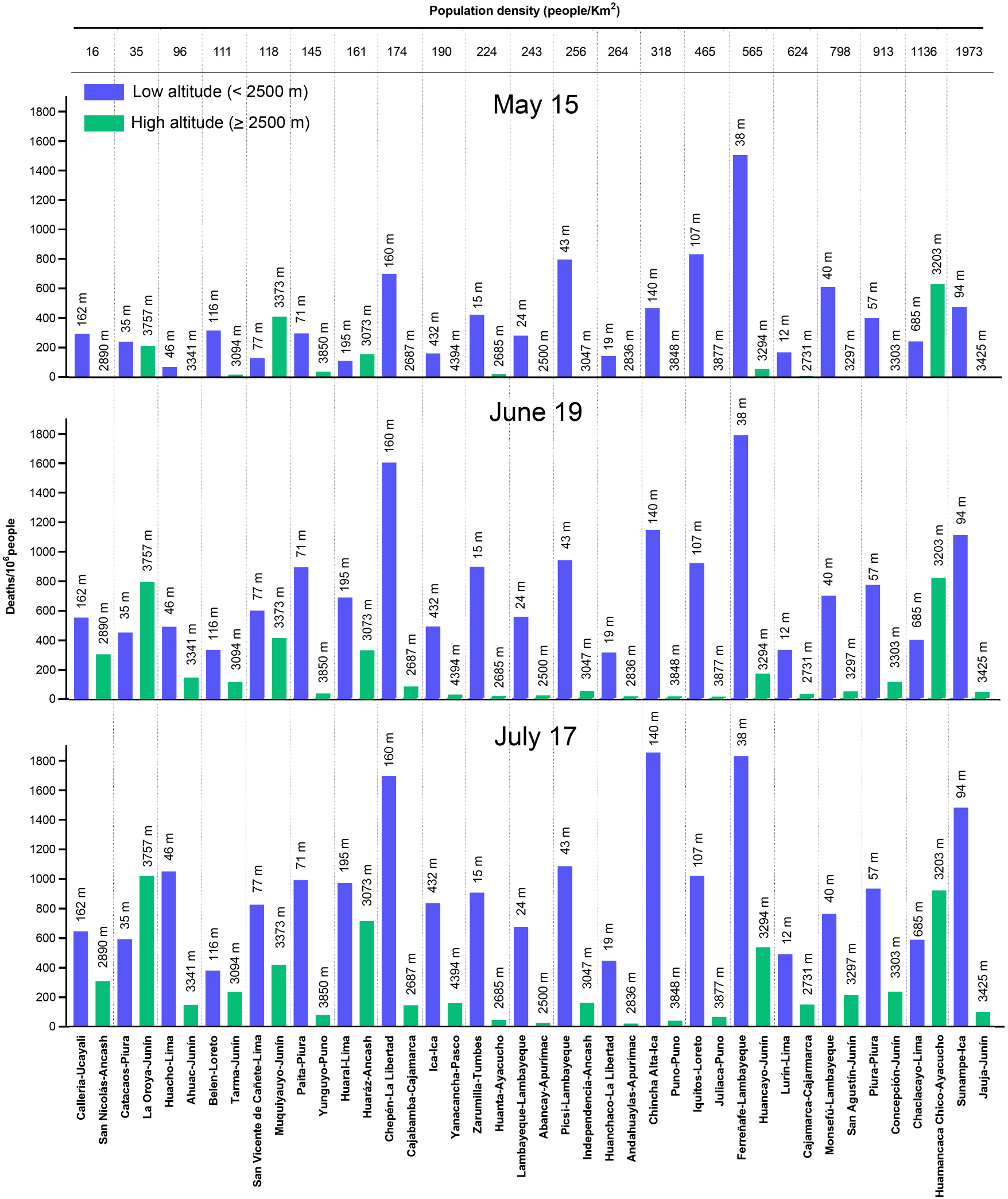
Pairwise comparisons for cumulative death rates between districts with equivalent population densities located at high or low altitudes. Death rates were compared in paired low-(< 2,500 m) and high-altitude (≥ 2,500 m) districts of similar population density (5% maximum difference). Comparisons were made at cut-off points May 15, June 19, and July 17.

### High incidence of COVID-19 at high altitude sites may reflect recent migration triggered by the COVID-19 pandemic

The above pairwise comparisons between districts with equal population densities and very different altitudes were plotted as fractions of the combined death rates of the two districts under consideration, for the May 15, June 19 and July 17 cut-off dates (Fig. 8). The earlier date showed a strong and general predominance of death rates in lower districts (< 2,500 m) over higher altitude districts (≥ 2,500 m), with two notable exceptions in the pairwise comparisons between the low-population density (35/Km^2^) districts Catacaos (Piura, low altituve) *vs*. La Oroya (Junín, high altitude) and the high-population density (1,136/Km^2^) districts Chaclacayo (Lima, low altitude) *vs*. Huamancaca Chico (Ayacucho, high altitude) (Fig. 8). This predominance was progressively attenuated on later cut-off dates in the comparisons Belén (Loreto, low altitude) *vs*. Tarna (Junín, high altitude), with a population density of 111/Km^2^, San Vicente de Cañete (Lima, low altitude) *vs*. Muquiyauyo (Junín, high altitude), with a density of 118/Km^2^, and Huaral (Lima, low altitude) *vs*. Huaraz (Ancash, high altitude), with a density of 161/Km^2^. This could be attributed to either decreased death rates over time in low-altitude districts or, conversely, increased death rates over time in the population density-paired high-altitude districts.

**Figure 8.**
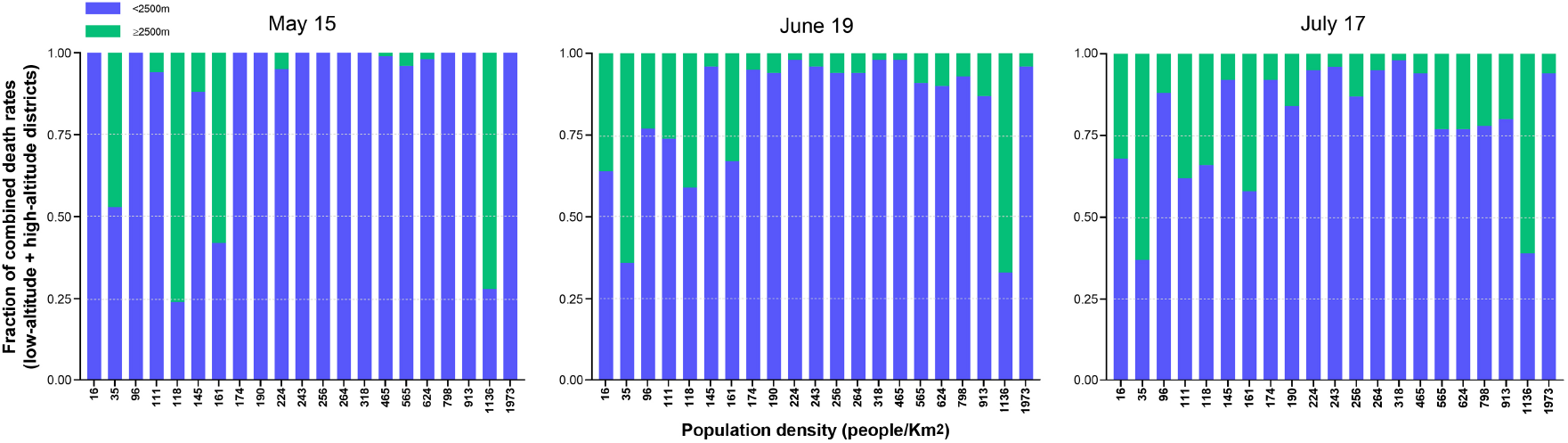
Graphical representation of fractions of combined death rates of population density-paired low-altitude *vs*. high-altitude districts. To illustrate the contribution of low and high-altitude districts of each pair, the fractional composition for all pairwise comparisons in Figure 6 was calculated as the number of deaths per million of each district, divided by the total number of deaths at each population density category.

More generally, we noted that, although high altitude sites had attenuated case and death rates as compared to low altitude sites, a number of high-altitude districts showed increased COVID-19 incidence and death rates over time. As an approach to trace the origin of these high-death rate districts located at high altitude, we plotted all districts as a function of deaths newly registered at three time intervals, from March 1 through April 10, April 11 through May 15, and May 16 through June 19. This showed that newly registered deaths from COVID-19 increased over time in a number of districts that clustered at altitudes between 2,500 and 3,500 m (Fig. S2). We reasoned that, under community spread, newly registered death rates would be in direct relationship to prior case rates in the same districts and thus be geographically distributed in accordance with prior case rates, while death rates associated with imported cases would not be correlated to prior local case buildup. Out of 743 districts in our database located at altitudes between 2,500 and 3,500 m, 65 had ≥ 50/10^6^ newly registered deaths between May 16 and June 19. Remarkably, geographical mapping of these districts revealed that 60 of them (92.3%) were located within a 50 Km-wide strip on either side of a path leading from the Lima metropolitan area through the Canta and Yauli districts and then along route 3N/3S, from the Cajamarca department in the North all the way to the Chilean border in the South, with a dense cluster near Huancayo, on route 3S (Fig. 9). Three of the 5 districts outside this path formed a tight cluster near Arequipa, a major urban center with high case and death rates. Interestingly, high-death rate, high-altitude districts first (April 11 through May 15 interval) appeared near Lima, and in more distant districts along the path described above on later dates (May 16 through June 19). Furthermore, all of the districts described above (Fig. 8) showing longitudinal shifts in death rate predominance in population density-paired comparisons between high- *vs*. low-altitude districts are located within this narrow strip of land. This pattern is highly suggestive of propagation from the early epicenter of the pandemic, namely the Lima metropolitan area, to high-altitude districts, which may explain the high-altitude, high-death rate clusters located in relative proximity to Lima and along a heavily transited path used by recent migration forced by the pandemic, in contravention of Government-mandated mobility restrictions.

**Figure 9.**
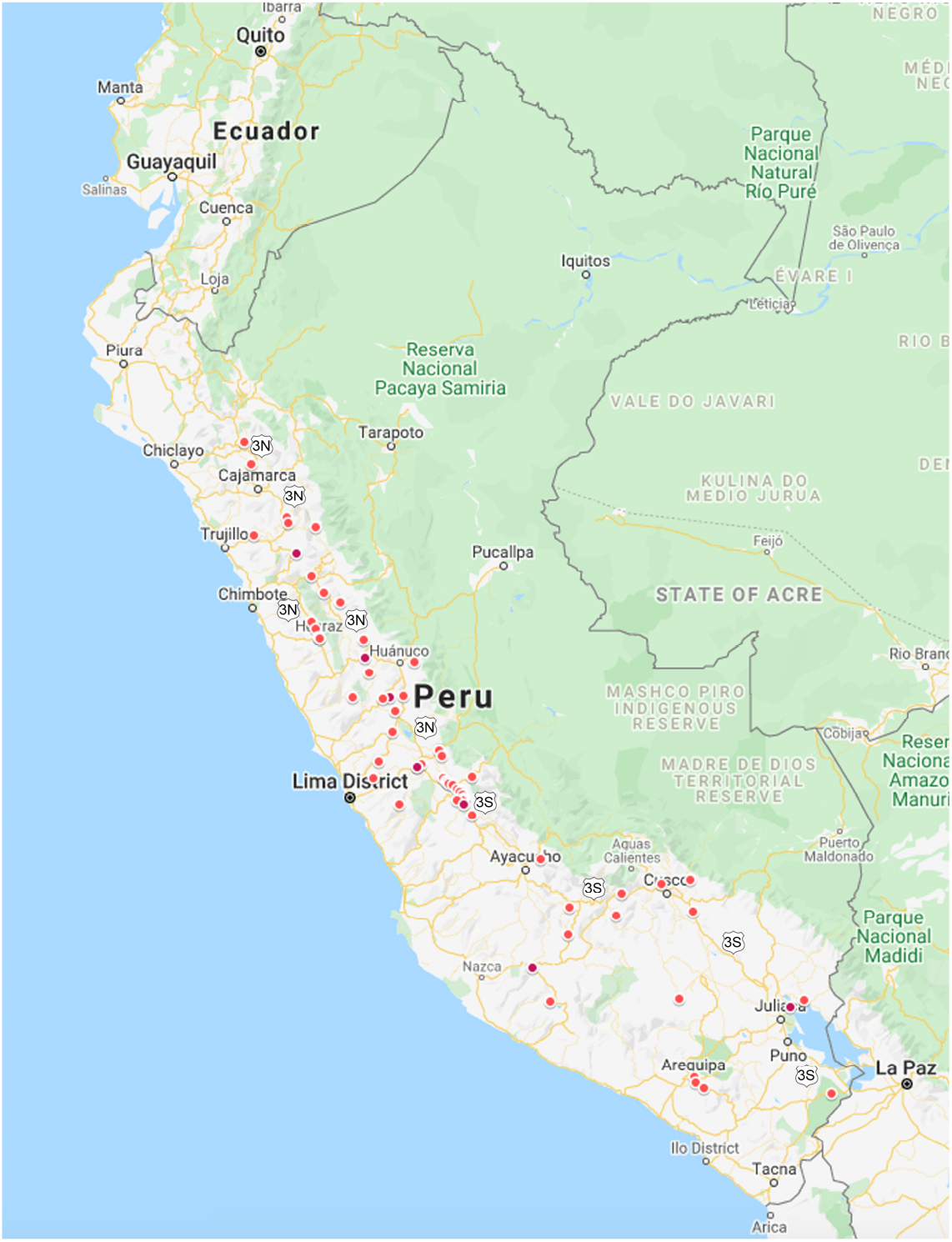
Geographical mapping of high-altitude districts with high death rates. All districts above 2,500 m of altitude with registered death rates ≥ 50/10^6^ people between May 16 and June 19, 2020 were mapped with the GoogleMyMaps online application. Highway 3N/3S is indicated at various points on the map. An interactive map can be accessed at https://www.google.com/maps/d/edit?mid=1VVUxlS9bz0rj-ChtbpGTQ0tUSs27TKWL&usp=sharing.

### Altitude as an independent protective factor from COVID-19

The above observations suggest that altitudes above 1,000 m confer protection from COVID-19. While the only known protective factor from the severity of this disease is young age^6-8^, several co-morbidities have been shown, in cohort studies, to associate with severe COVID-19, including hypertension, cardiovascular disease, obesity, diabetes or respiratory diseases^6-8,33^. The data described above (Fig. 1) hinted at several geographical differences in age distribution and male-to-female ratios of cases and deaths. To explore if age or sex are a factor associated with the diminished incidence and death rates in high-altitude districts, we compared the age distribution of COVID-19 cases and deaths in districts stratified by 1,000-m altitude intervals. There were no statistical differences in normalized case or death rate age distributions between altitude intervals on any of the four cut-off dates under consideration. A trend towards a younger age distribution in cases in the 3000-4999 m altitude range (Fig. 10A) was not statistically significant. Similarly, there were no significant differences in male-to-female case or death ratios in comparisons between different altitude intervals (Fig. 10B), although female death rates reached 0.50 at the 1000-1999 m range in the May 15 registry cut-off. From this, we conclude that neither altitude-dependent age distribution, nor differential sex susceptibility, explains the apparent protection from COVID-19 associated with high altitude.

**Figure 10.**
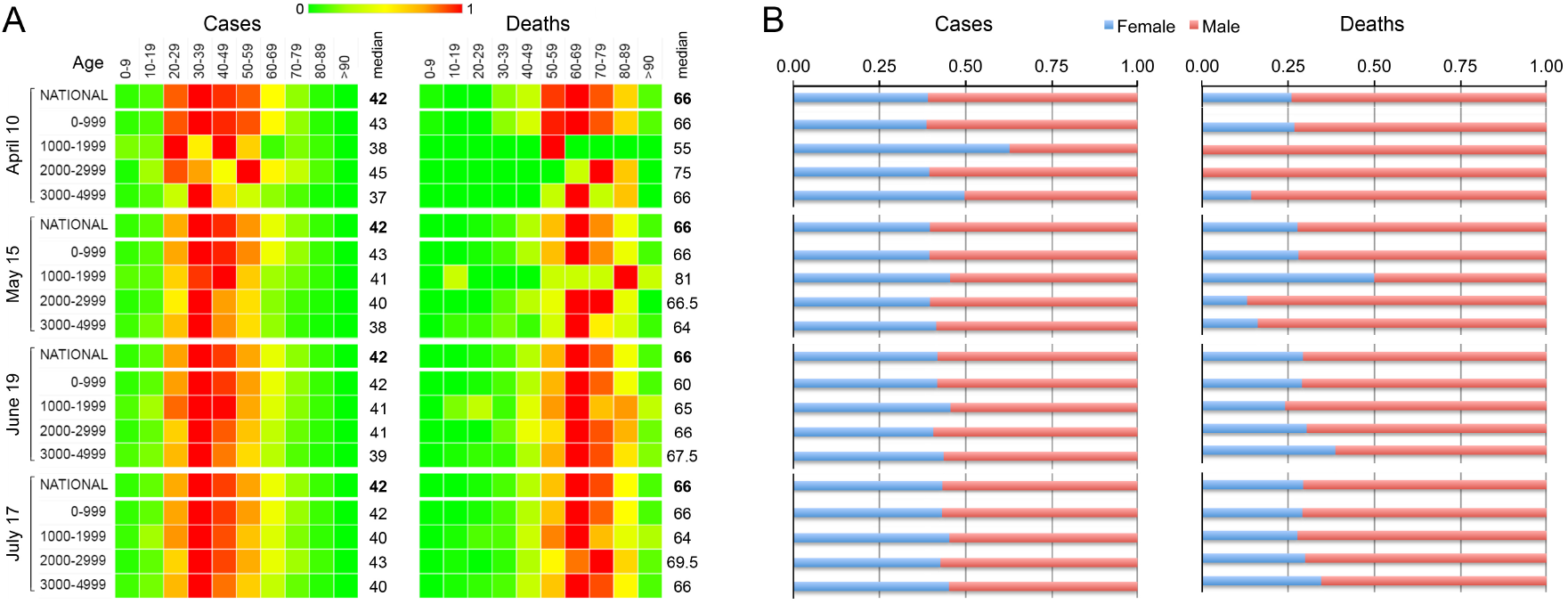
Age and sex distribution of COVID-19 cases and deaths in Perú as a function of 1000-m altitude intervals. **A**. Heatmap of frequencies of distributions of cumulative cases and deaths on each of the indicated dates in each of the indicated age ranges and altitude intervals. Median ages are shown to the right of the heatmaps. **B**. Histograms of female-to-male rates of cumulative and deaths on the indicated dates, and altitude intervals.

The dataset used in our analysis does not contain clinical information, which precludes the analysis of co-morbidities. In order to ascertain how co-morbidities known to associate with severe COVID-19 may influence the geographical distribution of cases and deaths, we resorted to a proxy approach by which the most recently available survey data on the nationwide prevalence of non-transmissible diseases in Peruvian cities^34^ were used as variables in principal component analysis (PCA) and correlation tests. The variables analyzed included prevalence of hypertension, hypercholesterolemia, diabetes, obesity, smoking habit and sedentary lifestyle, retrieved from the 2010 TORNASOL suvey^34^, the 2018 poverty index^35^, current population density, and the cut-off date of May 15 count for COVID-19 deaths per million people. This date was chosen assuming that it better reflects deaths from local cases than later dates, more subject to the influence of imported cases. Expectedly, our analyses showed clustering of sedentary lifestyle with obesity and hypercholesterolemia with diabetes (Fig. 11A). Interestingly, hypertension rates clustered more closely with COVID-19 death rates than with the other morbidities analyzed as variables. Rates of smoking habit did not cluster with any of the other morbidities, and population density was as close to COVID-19 death rates as were hypertension rates (Fig. 11A). Importantly, altitude inversely correlated with COVID-19 death rates and all morbidity prevalence rates analyzed. Also of note, poverty indexes clustered together with altitude, although without reaching statistical significance, and inversely correlated with all analyzed morbidities. (Fig.11B). With all the caveats stemming from a comparison between current epidemiological data and past prevalence of co-morbidities, plus the fact that we could only analyze select cities, these observations suggest that death from COVID-19 occurs preferentially in a context of high rates of morbidities such as hypertension and hypercholesterolemia, prevalent in low-altitude cities, and less preferentially in high-altitude, high-poverty index locations.

**Figure 11.**
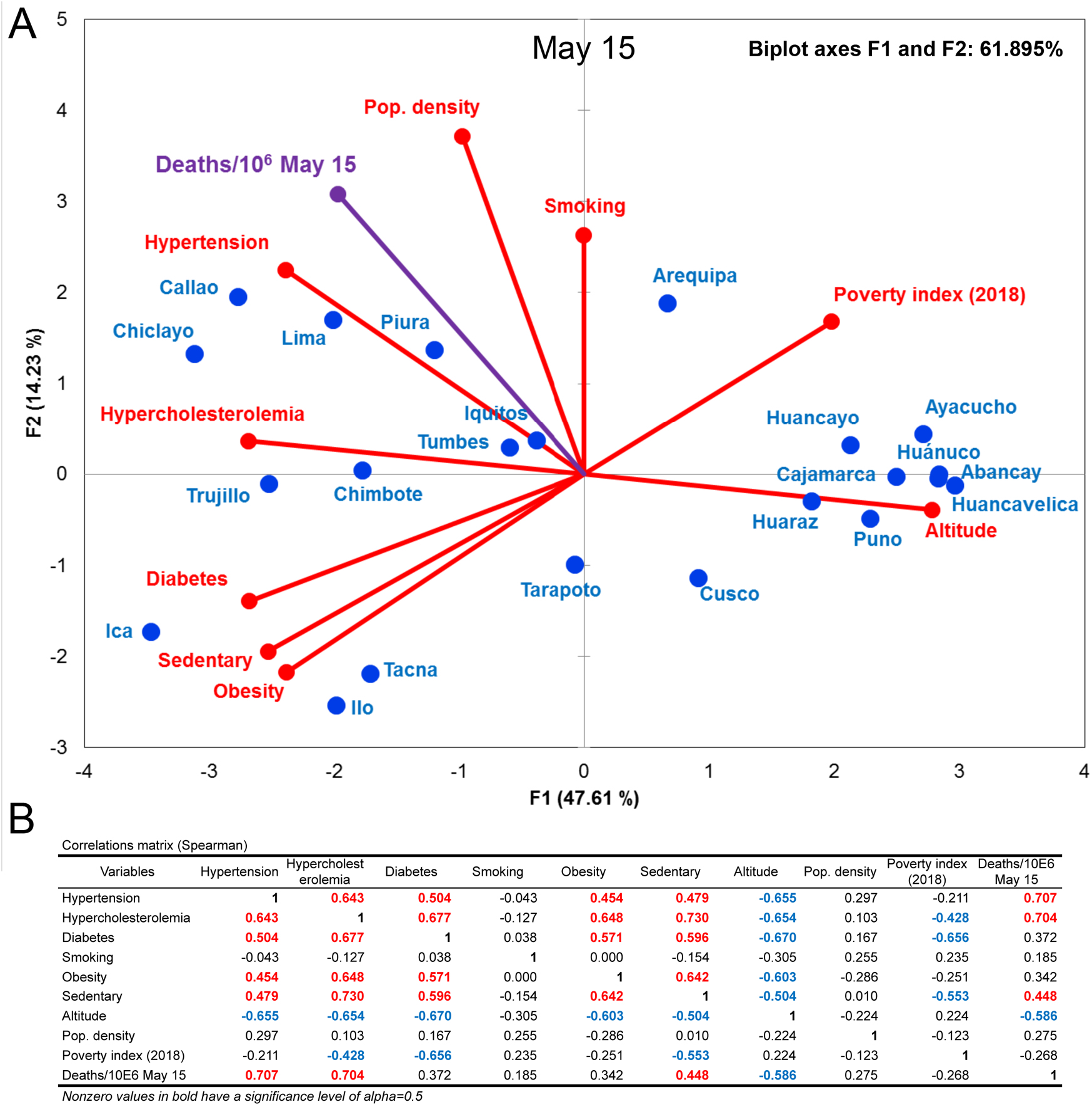
Correlation analyses of COVID-19 death rates, non-transmissible morbidity prevalence and altitude in major Peruvian cities. **A**. Principal component analysis of COVID-19 deaths registered from May 15 through June 19 (rates per 10^6^ population), together with morbidity prevalence, altitude and poverty index. **B**. Correlation analysis between variables. Values in bold are significant (alpha = 0.5; p ≤ 0.05). In red, positive correlation. In blue, negative correlation. P values for each correlation factor are provided in Table S5.

## Discussion

This ecological study is intended to provide details relevant for the understanding of the dynamics of the COVID-19 pandemic in Peru that may aid in shaping policy decision-making in the face of its unrelenting spread. Our most relevant finding is a significant association between life at high altitude and a lower risk of COVID-19 contagion and death from the disease. We also provide evidence that the recent surge of the pandemic in high-altitude districts in Peru may be explained by the influx of migrants from large urban centers to high-altitude districts along a specific land route, overriding the protective effect that might have been conferred by high altitude to residents of those particular districts.

Although prior studies have suggested that altitude may be a protective factor from COVID-19 (ref. 27,30-32,36-40), our analysis is the first to employ a large longitudinal database encompassing all COVID-19 cases and deaths registered in Peru through July 17, 2020 (https://www.datosabiertos.gob.pe/group/datos-abiertos-de-covid-19). In order to correct for population density, which tends to be significantly higher on the coastal lowlands, we have normalized case and death counts by population density and per million people in all districts. Likewise, we have considered the prevalence of non-transmissible diseases as determined by prior surveys, as a proxy for potential co-morbidities associated with case and death incidence^34^. Past studies have shown that low-altitude populations in Peru, both on the coast and in the Amazon forest departments, have significantly higher risk than high altitude populations for a number of non-transmissible diseases, including obesity, diabetes or hypertension^41,42^, comorbidities that impart strong risks for the development of severe COVID-19 (ref. 6-8,33).

Our multivariate analysis corroborates that the prevalence of obesity, diabetes, hypertension and hypercholesterolemia correlate with low altitude. It further reveals that the normalized death rate from COVID-19 is significantly correlated with prevalence of hypertension and hypercholesterolemia, as well as low altitude. Interestingly, death rates from COVID-19 are not significantly correlated with population density, despite a trend towards correlation. Indeed, our comparisons of case and death incidence between populations stratified by altitude shows the same protective effect of altitude when normalized by million people or by population density (population/Km^2^). Although crowding is a known viral spreading factor^43^, there is evidence that spread of SARS-CoV-2 tends to adopt focal patterns, with superspreader conditions or individuals in which high viral production and loads can be more relevant nucleating transmission factors than mere population density^43-45^. Likewise, current COVID-19 death rates are not correlated with local prevalence of smoking habit, an observation consistent with a recent large-scale cohort study that found that smoking did not confer a significantly increased risk of severe COVID-19 (ref. 8).

It could be argued that populations in sparsely populated sites at high altitude might have less access to diagnostic PCR or serological tests confirmatory of SARS-CoV-2 infection. However, these tests are applied to the general public free of charge by the Peruvian national health system, and there is no evidence of shortage of serological tests, although they are unevenly distributed throughout the territory (see Study Limitations). Likewise, high poverty levels could limit the availability of health care and may be expected to be associated with elevated case and death rates as observed elsewhere (e.g., https://www1.nyc.gov/site/doh/covid/covid-19-data.page). Contrary to this expectation, in our multivariate analyses, high poverty indexes were associated with higher altitude and lower, not higher, death rates. In conclusion, altitude appears to confer protection from COVID-19 incidence and death independent of population density and not offset by high poverty levels. The latter observation is at odds with the findings of the OpenSAFELY study, which detected an increased risk of COVID-19 death with greater deprivation in the United Kingdom^8^, not fully explained by pre-existing conditions or other risk factors. This reinforces the notion that additional local conditions are significant modifiers of sensitivity to COVID-19.

We have found that the apparent protective effect of altitude was very strong when comparing populations below and above the accepted high-altitude threshold (2,500 m), with relative risks for low-altitude populations, both in coastal and Amazonian regions, between 6.5 and over 11-fold for cases and over 6.5 and 21.6-fold for deaths, relative to high-altitude populations. Because atmospheric pressure decreases with altitude, inspired PO_2_ (PO_2_) is also reduced. At 2,500 m, PiO_2_ decreases approximately to 110 mm Hg from its sea-level value of 150 mm Hg, and consequently arterial PO_2_ drops below 70 mmHg. Around this value, hemoglobin (Hb) O_2_ saturation approaches the inflexion region of the O_2_-Hb dissociation curve and therefore the physiological effects of hypoxemia become noticeable and significant. Hence the use of 2,500m as the threshold for defining high altitude. Our finding that population density-adjusted COVID-19 death rates gradually decline from altitudes below 500 m up to 2,000 m is suggestive of an effect of chronic high-altitude hypoxia exposure associated with the seeming protective effect from COVID-19 conferred by altitude.

In spite of the general potential protection conferred by altitude, we have observed clusters of high incidence of cases and deaths from COVID-19 at high-altitude sites. Geographical localization of high-altitude clusters of newly registered COVID-19 deaths revealed that the majority corresponds to sites on, or adjacent to, a major migration route from the Lima metropolitan area, namely route 3N/3S. Because the overwhelming majority of the high altitude locations outside of this route have very low rates of COVID-19 cases and deaths, we surmise that these high case and death rate clusters are a reflection of viral spread imported from the high-rate Lima metropolitan district, rather than *bona fide* community spread. In all transmissible disease epidemics, short distance from a major focus is a predictor of case incidence and rate. However, in the COVID-19 pandemic in Peru, sites located at long distances from Lima, such as districts in the Amazon regions, show high case and death rates. Importantly, the latter sites are located at low altitude. Based on these considerations, we suggest a scenario in which altitudes above 1,000 m confer a general protective effect from COVID-19, in co-existence with high case and death rate clusters imported from Lima. At this time, we have not observed evidences for spread to high-altitude sites from other high-incidence coastal sites, a situation that might change if the pandemic continues its current trend.

This scenario implies that the apparent protection from COVID-19 conferred by altitude would benefit long-time resident populations, rather than recent migrants to high-altitude districts. Protection attributed to altitude was also noted during the 2009 H1N1 influenza pandemic in Peru^46^ and the 2006 H5N1 avian influenza epidemic in Indonesia^47^ and Thailand^48^. Experiments with mice acclimatized to high altitude also showed enhanced resistance to a type A influenza strain^49^. As noted above, a major factor impacting physiological changes associated with altitude is hypobaric hypoxia^50^. Low environmental PO_2_ at high altitude is a selective force that has shaped the genetic endowment of highlanders in the Andes, the Himalayas and the Ethiopian plateau^51^. Human populations have evolved to select for local and shared high-altitude adaptive traits. Adaptive variants in regulators of the hypoxia response pathways are commonly shared among populations from all three high-altitude regions^51,52^, including EPAS1 (also known as HIF-2), SENP1 or EGLN1 (PHD2) in the Andes^53^ and the Himalayas^54^ or THRB and ARNT2 in Ethiopia^55^. The implication is that these highland populations display genetically-determined elevated steady-state activation of hypoxia-response pathways compared to lowlanders. This is supported by evidences such as differences in lung respiratory capacity, peripheral and central vasculogenesis, enhanced erythropoiesis and metabolic signatures^50^, all of which are known targets of the hypoxia response in cellular and animal models^56^. Some of these physiological responses are also observed in lowlanders upon acute hypoxia or under short-term acclimatization to high altitude, which are reversed at low altitude.

In keeping with our hypothesis that long-term, but not short-term exposure to altitude and relative hypoxia may explain protection from COVID-19, we propose that stable morphological, physiological, and metabolic characteristics resulting from long-term or life-long high altitude exposure, genetically or epigenetically determined, are more likely to be associated with relative protection from severe respiratory viral diseases than short-term responses to hypobaric hypoxia. Potential mechanistic explanations of the protective effect of altitude might include chronic hypoxia-induced adaptive characteristics in the lungs at the organ and cellular level, expression of viral receptors and associated endothelial and lymphatic vessel networks, as well as metabolic adaptations.

SARS-CoV-2 uses angiotensin-converting enzyme 2 (ACE2) for cell entry, which is potentiated by the transmembrane serine protease 2 (TMPRSS2)^57^. As such, low levels of both proteins could mitigate the entry of SARS-CoV-2 into susceptible cells. Hypoxia and the activation of hypoxia-dependent pathways have shown to modulate the expression of ACE2 and TMPRSS2. Acute hypoxia *in vitro* induces HIF-1-independent expression of ACE2^58,59^, followed by HIF-1-dependent repression and downregulation under prolonged hypoxia^60^. Also, activation of the HIF pathway through the hypoxia-mimicking compound CoCl_2_ causes a decrease of TMPRSS2 expression *in vitro*^61^. Downregulation of ACE2 by chronic hypoxia has also been proposed as a mechanism for pulmonary hypertension associated with acute mountain sickness in lowlanders travelling to high altitude^62^. However, there is no evidence that ACE2 expression is regulated *in vivo* in populations living at high altitude^63^. Furthermore, this possible mechanism would be specific of viruses using ACE2 as their receptor, such as SARS-CoV and SARS-CoV-2, and would not explain protection from H1N1 influenza or viruses that use other cellular ports of entry.

On the other hand, low ACE2 levels are expected to lead to angiotensin-II accumulation with consequent pulmonary vasoconstriction and vascular remodeling resulting in increased pulmonary artery pressure (PAP)^60,62,64^. The usual mild to moderate pulmonary hypertension observed in Andean highlanders^65^ is somewhat compensated by chronic hypoxia-induced lung vascularization preventing PAP to reach exaggerated values. In addition, augmented lymphangiogenesis may facilitate fluid drainage to prevent lung edema, as shown in some animal models^66^. Importantly, experimental genetic depletion of lymphatic vessels leads both to lung emphysema and lung fluid accumulation under injury^67^, highlighting the importance of the lymphatic network in normal and diseased lung function. Mechanistically, hypoxia induces HIF-1-independent expression of the lymphangiogenic factors VEGF-C and VEGF-D^68^, promoting new lymphatic vessel formation, possibly in concert with hypoxia-driven modulation of Notch activity^69^. Additionally, inflammatory stimuli^70^ and stretch injury^71^ stabilize HIF-1α in alveolar epithelial cells, and resolution and repair in the alveolus requires the proliferation and trans-differentiation of alveolar cells, that in turn requires HIF-1α-dependent VEGF expression^70^. It should be noted that the lung injuries associated with COVID-19 are distinct from those of high-altitude pulmonary edema (HAPE)^72,73^. While the acute respiratory distress syndrome in COVID-19 is due to primary alveolar and lung endothelial destruction with concomitant inflammation and microthromboses^74,75^, HAPE is a non-cardiogenic pulmonary edema caused by hypoxic pulmonary vasoconstriction that leads to a non-inflammatory and hemorrhagic alveolar capillary leak that only over time may elicit a secondary inflammatory response^76^. Nevertheless, it is plausible that the protective pathophysiological mechanisms discussed above are relevant to both conditions.

Of potential relevance in the context of COVID-19, hypoxia has significant consequences on the activity and regulation of the immune and inflammatory systems and metabolic networks. Like in SARS or MERS, in addition to acute respiratory distress and, less frequently, heart or renal failure^77^, uncontrolled hyperinflammatory states^78^ and disseminated coagulopathy^77,79^ underlie death from COVID-19 in many patients. Hypoxia results in metabolic and immune-inflammatory reprogramming and activation of HIF and NF-κB pathways in tightly intertwined networks^80^, and adaptation to high altitude and chronic hypoxia follows different metabolic and immune modulatory routes for lowlanders and highlanders^81^. In experimental settings, acute hypoxia favors a HIF-1α-mediated skewing of macrophages to an inflammatory M1 phenotype, and of immunosuppressor Treg to inflammatory Th17 phenotpes, dependent on increased mitochondrial succinate production. In contrast, HIF-2α polarizes an inflammatory-suppressing M2 macrophage phenotype^82^. Of note, while the initial response to hypoxia (< 24 h) is dominated by HIF-1α, more sustained responses are supported by HIF-2α^83^. Also relevant when considering immune and inflammatory adaptation to hypoxia and high altitude, HIF-1 is ubiquitously expressed, whereas HIF-2 shows more restricted expression patterns, predominantly in immune cell subtypes such as macrophages, neutrophils and lymphocytes^84^.

In a study of effects of short-term and long-term exposure to high altitude, lowlanders, but not Sherpas, showed diminished muscle succinate levels while developing signs of oxidative stress with prolonged exposure to high altitude, at least partly owing to lower but more efficient OXPHOS in altitude in Sherpas^81^. HIF-1α promotes glycolysis through transcriptional upregulation of glycolytic enzymes, while limiting mitochondrial respiration through upregulation of pyruvate dehydrogenase kinase-1 (PDK1), which inhibits pyruvate dehydrogenase and entry of pyruvate to the TCA cycle^56^. Consistently, in the same study, lowlanders showed increased glycolytic activity at high altitude, while Sherpas had no sign of altered glucose homeostasis^81^, suggesting that highlanders have reprogrammed their metabolism for alternative sources of ATP in hypoxia. As high glycolytic rates favor the differentiation of myeloid progenitors to proinflammatory M1 macrophages^80^, this would provide a mechanistic explanation for the inflammatory reaction observed in lowlanders at high altitude and the protection form excessive inflammation in highlanders. In turn, blunted inflammatory responses are expected to protect highlanders from inflammation-induced obesity and adipocyte accumulation, another source of inflammatory molecules such as interleukin-1β (IL-1β) or interleukin-6 (IL-6)^85,86^. Interestingly, apart from HIF-pathway genes, variants coding for metabolic and immune-inflammatory molecules are significantly linked with highlander populations, including IL1B, IL6 and tumor-necrosis factor (TNF) among Andeans and Himalayans, or peroxisome proliferator-activated receptor alpha (PPARA) among Himalayans^51^.

Finally, erythropoietin (Epo) is also a candidate to play an anti-inflammatory role in highlanders following SARS-CoV-2 infection^87^. Besides its role in erythropoiesis, this pleiotropic hormone/cytokine exerts neuro and immunomodulatory, anti-inflammatory, anti-apoptotic, and tissue-protective effects^88,89^, and thus could be beneficial in diminishing the clinical outcome severity and contribute to explain in part the lower lethality due to COVID-19 at high altitude. Chronic hypoxia increases Epo expression in most tissues and reduces the expression of the soluble Epo receptor (sEpoR), an endogenous Epo antagonist^90,91^. This combination increases Epo availability and ensures a stronger Epo signaling. Andean highlanders show modestly increased serum Epo and decreased sEpoR compared to lowlanders^92^, and therefore it is possible that an Epo-related anti-inflammatory steady-state favors a better clinical outcome after SARS-CoV-2 infection.

In summary, we favor the view that long-term high altitude exposure protects from severe respiratory viral diseases, including COVID-19, through multiple mechanisms prevalent in residents or genetically selected populations adapted to chronic hypobaric hypoxia. Such mechanisms include physiological, anatomical, metabolic and immune-inflammatory adaptations. The role played by unabated inflammation in COVID-19 pathogenesis has been evident from the outset of the pandemic^78^ and addressed therapeutically by targeting inflammatory molecules, and that of metabolic responses and viral subversion has been eloquently highlighted in a recent review article^93^. It has been argued that environmental factors, including high levels of ultraviolet radiation, low humidity or low temperatures, could contribute to a diminished transmission of SARS-CoV-2 at high altitude^63^. It should be noted, though, that high humidity acts as a protective barrier against^94,95^, while low temperatures favor^95^, viral respiratory diseases. Our analysis supports that living in altitude is a general protective factor from severe COVID-19. Translating this knowledge to therapies for immediate use against COVID-19, for instance by means of pharmacological stabilizers of HIF transcription factors^93,96^, is challenging, given the likelihood that protective effects may require long-term multi-systemic adaptations and that adroit navigation will be required to negotiate through the heterogeneous downstream consequences of manipulating different components of the hypoxia response.

## Study limitations

This study presents some limitations. First, the public databases used for our analyses contain gaps and inconsistencies in the numbers of cases or deaths reported for some districts. As described in the Methods section, all such districts were removed from our analyses, which might introduce a bias. The removed districts, though, showed geographical distributions that were similar to the entire set of districts, particularly with regards to altitude. As such, removal of these districts from our analysis is not expected to result in significant variations in our conclusions pertaining to the associations between COVID-19 incidence and altitude.

Second, positive cases encompass those tested by more robust (sensitive, specific and reproducible) qPCR diagnostic assays and those tested by arguably less reliable serological assays^97^. The latter may include false positive results, potentially affecting our analysis and conclusions. While considering this issue as potentially problematic, we have also reasoned that such a bias would be similarly represented in all districts, thus largely offsetting possible distortions introduced by false positives in our comparative analyses between different districts or groups of districts.

Third, although data on testing coverage per district or province (encompassing several districts) are not available, data per department (the next level of territorial organization), and show variations in coverage from 1.08% of the population in Cajamarca to 8.76% in Moquegua (Table S3). *A priori*, this could introduce a bias towards underreporting in registered cases below uncertain thresholds of testing coverage, while testing coverage above yet uncertain thresholds would result in lower rates of registered positive cases. In general, larger coastal urban centers have higher testing coverage. Systematic population-level (> 5,000 individuals) serological testing for SARS-CoV-2 has found case prevalence between 1.7% and 19.3% in diverse populations with very different testing coverage^98-100^. However, we have found no significant correlation between testing coverage and case rates (R^2^ = 0.13) or, particularly, death rates (R^2^ = 0.03) (Fig. S3). Accordingly, we have placed more emphasis on the conclusions drawn from the analysis of death rates.

Fourth, our correlation analyses of COVID-19 deaths with the prevalence of non-transmissible diseases uses, for the latter, data from a survey conducted in 2010-11 in select cities in Peru. More recent surveys^101,102^ provide information limited to one or two the morbidities of our interest and without city- or district-level detail. Although the relative prevalence among departments of hypertension, obesity or smoking habits did not vary significantly between 2014 and 2019 (ref. 102), our analyses would provide a more accurate representation with more current prevalence data for morbidities at city or district levels.

Finally, this study, like many other current studies on the COVID-19 pandemic, suffers from the hurdle posed by a constantly moving target. As such, our conclusions on the protective effects of altitude, supported by our correlation, longitudinal and geographical analyses, may risk being quickly obscured *de facto* by the massive and rapid pandemic wave currently sweeping the entire Peruvian territory. Imported cases quickly turn into foci of community transmission, with potential high viral loads overwhelming any possible protective effect that may be afforded by physiological or molecular adaptations to high altitude of the affected local populations. In other words, adaptations to high altitude may confer a degree of protection from, but not immunity against, SARS-CoV-2 and its pathogenicity.

## Methods

### Study design

We conducted a retrospective study using national registry data including COVID-19 cases and death data (see Data Source). The study explores cumulative data registered on four different cut-off dates: April 10, May 15 and June 19, July 17. Cases correspond to the population diagnosed as infected with SARS-COV-2 and positive by either quantitative RT-PCR (molecular test) or by a serological assay. Deaths from COVID-19 correspond to patients positive for SARS-CoV-2 by either a molecular or a serological assay, and certified as death from COVID-19.

### Data source

Data on positive COVID-19 cases and deaths were collected from the Open COVID-19 data resource released by the Secretaría de Gobierno Digital of the Peruvian Government. Data was accessed between June 10, 2020 and July 17, 2020 through https://www.datosabiertos.gob.pe/group/datos-abiertos-de-covid-19. The data file provided by the Open Data contains country-specific daily numbers of COVID-19 cases and deaths, and include gender, age, date of diagnosis, type of diagnostic assay (serological or PCR), and location information including district, province and department. For each date, we calculated two COVID-19 epidemic measures: i) number of cases per million people or population density calculated using the number of cumulative cases and population size or population density, ii) number of deaths per million people or population density calculated as the number of cumulative deaths and population size or population density. Demographic data for Peru was obtained from the Instituto Nacional de Estadística e Informática (INEI) through https://www.inei.gob.pe. Data were a cumulative count up to and including April 10, 2020; May 15, 2020, June 19, 2020, and July 17, 2020. Records were considered as nulls when age, gender or date were missing and when district and province were missing. The resulting dataset includes 333,958 cases and 12,045 deaths as of July 17, 2020. For some analyses, cases and deaths newly registered between April 11 and May 15 or between May 16 and June 19 were retrieved. Each case represented a positive diagnosis of COVID-19 along with demographic data.

### Demographic parameters

A descriptive analysis was performed on the basis of nationwide data for gender and age of cases and deaths, stratified into 10 age ranges (0-9, 10-19, 20-29, 30-39, 40-49, 50-59, 60-69, 70-79, 80-89, ≥ 90). Data was analyzed both by departments and by altitude intervals (in meters above sea level), the latter associated with each district. Departments included Amazonas, Áncash, Apurímac, Arequipa, Ayacucho, Cajamarca, Callao (Constitutional province), Cusco, Huancavelica, Huánuco, Ica, Junín, La Libertad, Lambayeque, Lima, Loreto, Madre De Dios, Moquegua, Pasco, Piura, Puno, San Martín, Tacna, Tumbes y Ucayali. Districts were classified according to their altitude in the following ranges: 0-999 m, 1000-1999 m, 2000-2999 m, and 3000-4999 m. The altitude for each district as well as data on population size, territory area, and population density were obtained from the National Institute of Statistics and Informatics (INEI) at available at https://www.inei.gob.pe/. This database is available in Table S1.

### Mapping

Geographical maps with cases and deaths per districts were composed with Mapinseconds (http://www.mapinseconds.com/) and precise geographical localization of selected districts was done with the aid of GoogleMyMaps (https://www.google.com/mymaps).

### Estimates of Relative Risk

The relative risk (RR), its standard error and 95% confidence interval were calculated according to Altman^103^. The RR is given by

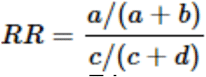

with the standard error of the log relative risk being

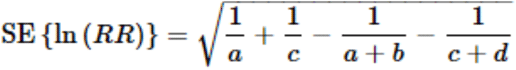

and the 95% confidence interval

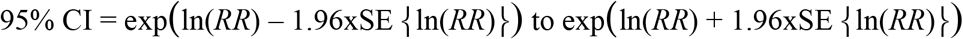

RRs were calculated for pairwise comparisons of cumulative cases or deaths between different altitude intervals on a given cut-off date. Cases or deaths counts were designated events (numerators), and the sums of the general population for each altitude interval were designated controls (denominators). Population numbers as control values was used rather than numbers of SARS-CoV-2 diagnostic tests performed, given the uncertainty and the territorial unevenness of the testing coverage.

### Pairwise comparisons of low- and high-altitude districts

Official reports show total number of deaths by department. However, many of these regions harbor low- and high-altitude provinces. Thus, to exemplify the proportional contribution to total death rates of these provinces, we compared death rates normalized to population density of low and high-altitude capital districts in the same department. To illustrate the relationship between death rate, altitude and population density, we compared death rates in paired low- and high-altitude districts of similar population density. Pairs of districts were generated based on their altitude (< 2,500 m or ≥ 2,500 m) and a maximum population density difference of 5%. Comparisons were made at cut-off points May 15, June 19, and July 17. To illustrate the contribution of low and high-altitude districts of each pair, the fractional composition was calculated as the number of deaths per million of each district, divided by the total number of deaths for each population density category.

### Principal component analysis (PCA) and correlation tests (CT)

Cities were selected for which prevalence data for morbidities are available as part of the TORNASOL survey^34^. For the same cities, the 2018 poverty survey data^35^ were used, and COVID-19 death rates (per 10^6^ people) on the May 15 cut-off date were retrieved from the curated general dataset. Altitude and population density were retrieved as indicated under Data Sources. Values for all variables were row median-normalized and subjected to PCA and CT as implemented in XLSTAT. For graphical representation, the first two factors from PCA were plotted as biplots including observations and variables with their depicted eigenvectors.

### Generation of heatmaps

For heatmap generation, variables were row-normalized and heatmaps generated with the aid of Morpheus (https://software.broadinstitute.org/morpheus/).

### Other statistical analyses

For non-paired, non-parametric data comparisons, Mann-Whitney tests were performed on GraphPad. Chi-square tests were applied for contingency data analyses. Unless otherwise indicated, significance levels are denoted in graphs by asterisks, as follows: * ≤ 0.05; ** ≤ 0.01; *** ≤ 0.001; **** ≤ 0.0001.

## Data Availability

All data referred to in the manuscript are available at the URLs below.

https://www.inei.gob.pe/media/MenuRecursivo/publicaciones_digitales/Est/Lib1526/index.html.

https://www.inei.gob.pe/media/MenuRecursivo/publicaciones_digitales/Est/Lib1718/Libro.pdf

https://www.datosabiertos.gob.pe/group/datos-abiertos-de-covid-19

https://covid19.minsa.gob.pe/sala_situacional.asp

https://cs.ucsp.edu.pe/monitor-covid/

## Acknowledgments and Funding Sources

This study was partially funded by grants from the CSIC (COV19-006) and CIBER-EHD (Spain), awarded to T.M.T. F.C.V is funded by a Wellcome Trust Intermediate Research Fellowship in Public Health and Tropical Medicine (107544/Z/15/Z).

## Author Contributions

C.M., T.M.T. and F.C.V. conceived and designed the study, analyzed the data, generated figures and wrote and assembled the manuscript; H.A.G., F.C. and R.J.F. retrieved and curated databases, analyzed data and generated figures.

